# Estimation and monitoring of COVID-19’s transmissibility from publicly available data

**DOI:** 10.1101/2020.05.24.20112128

**Authors:** Antonio Silveira, Antonio Pereira

## Abstract

The COVID-19 pandemic began in the city of Wuhan, China, at the end of 2019 and quickly spread worldwide. The disease is caused by contact with the SARS-CoV-2 virus, which probably jumped from an animal host to humans. SARS-CoV-2 infects various tissues in the body, notably the lungs, and patients usually die from respiratory complications. Mathematical models of the disease have been instrumental to guide the implementation of mitigation strategies aimed at slowing the spread of the disease. One of the key parameters of mathematical models is the basic reproduction ratio *R*_0_, which measures the degree of infectivity of affected individuals. The goal of mitigation is to reduce *R*_0_ as close or below 1 as possible, as it means that new infections are in decline. In the present work, we use the recursive least-squares algorithm to establish the stochastic variability of a time-varying *R*_0_(*t*) from eight different countries: Argentina, Belgium, Brazil, Germany, Italy, New Zealand, Spain, and the United States of America. The proposed system can be implemented as an online tracking application providing information about the dynamics of the pandemic to health officials and the public at large.

## 1 INTRODUCTION

On the 11th of March 2020, the World Health Organization (WHO) declared the 2019 coronavirus disease (COVID-19) a global pandemic [1]. COVID-19 is caused by the SARS-CoV-2 coronavirus and was first reported in Wuhan, China, in December 2019 [2]. Since then, COVID-19 has spread globally with millions of laboratory-confirmed cases and hundreds of thousands of deaths [3]. The median incubation period of COVID-19 is 5.1 days and nearly all infected persons who have symptoms will do so within 12 days of infection [4]. However, an unprecedented characteristic of COVID-19 is its capacity for asymptomatic transmission [5], which contributes to increase the probability of transmission [6]. So far, there is no specific treatment for the disease and many research teams are currently working on a vaccine that, optimistically, will only be available in 2021. Even then, it will take some time to inoculate a significant share of the population. Meanwhile, hospital structures around the globe (e.g. intensive care units’ (ICU) beds, ventilators, etc.) are becoming overwhelmed with new patients and the increasing caseload will prove most catastrophic for poor countries, which lack the adequate health-care capacity to deal with the unparalleled demand posed by COVID-19 [7, 8].

So far, efforts to contain the spread of the disease have focused on the adoption of non-pharmaceutical interventions (NPI) based on population behavioral change and social distancing, such as banning large gatherings, enforcing the use of face masks, washing hands, or imposing severe lockdowns [9, 10]. Due to significant uncertainties regarding the transmissibility of SARS-CoV-2 as well as other political, social, and economic considerations, it is necessary to delineate effective social distancing policies which are able to alleviate COVID-19’s burden on healthcare structure and borrow time for the development of a vaccine or drug candidates while also simultaneously reducing the socioeconomic strain of living in a locked-down, confined society [11]. The effective monitoring of the epidemic’s dynamics plays a crucial role in the ongoing containment efforts, but will also continue to do so for some time ahead when social distancing control measures are eventually relaxed and the first wave of the pandemic is followed some months later by second or third waves of infection that may be more severe than the first [12].

Mathematical models, by providing a quantitative framework for hypothesis evaluation and the estimation of changes in transmission of infectious diseases over time and space, can indicate whether containment measures are having a measurable effect while guiding the design of alternative interventions [13]. Mathematical models vary in many aspects, including complexity in terms of the number of variables and parameters used, spatial and temporal resolution (e.g., discrete vs. continuous-time), and design ‘ (e.g., deterministic or stochastic) [14]. Mechanistic models of the susceptible-infected-recovered SIR type [15, 16] are the standard framework for a wide array of infectious diseases, including COVID-19 (see,) for example, Li et al. [17], Weitz et al. [18]). However, parameter estimates for a given model are subject to two major sources of uncertainty: the noise intrinsic to the data and the ad-hoc assumptions used for ascertaining parameter estimates [14].

The basic reproduction number [19], *R*_0_, or the average number of new infections caused by an infectious individual [20], is widely used to characterize the dynamics of infectious outbreaks and guide policy-planning. For instance, the average *R*_0_ at the start of the SARS pandemic in 2003 was estimated to be around 2.75 and was later reduced to 1 due to intervention strategies, including isolation and quarantine activities [21]. *R*_0_ is an imprecise estimate that is rarely measured directly (it is the product of disease parameters: duration, opportunity, transmission probability and susceptibility - DOTS) and rests on particular model structures and assumptions [22]. Modellers face many challenges when trying to provide robust estimations) of *R*_0_ in the current pandemics, such as the existence of superspreaders, the fact that SARS-CoV-2 can also be spread by asymptomatic individuals, and the scarce availability of testing supplies [17]. Several methods have been proposed to track trends in *R*_0_ during the course of an epidemic [23, 24, 25, 26, 27]. The access to reliable estimates of *R*_0_ could provide useful information about the efficacy of containment measures and allow their effective management in order to keep hospitalization rates within a desired approximate range [28].

In the present study, we model the transmission dynamic of COVID-19 in eight countries (Fig. 1) using least-squares algorithm (LSA) techniques. The criteria used to select the eight countries were due to their ‘ geographical location in either the northern/southern hemisphere or due to specificities regarding their first response to the pandemic threat as reported in the news media. Some (e.g. New Zealand, Argentina) were l very effective in responding rapidly to the pandemic by implementing lock down measures, while others (e.g. USA and Brazil) initially downplayed the virus’ threat and took longer to initiate containment protocols. Our goal is to contribute to understanding the spread of SARS-CoV-2 and compare *R*_0_ uncertainty arising from noise in the time series data gathered from public online sources. We used machine learning algorithms to optimally estimate a time-varying *R*_0_(*t*), which allows the monitoring of the ongoing pandemic in almost real-time. We compared the daily country reports for *R*_0_ to those we estimated in the present work in order to assess the reliability of official data. Besides, LSA-based techniques were used to reveal clues regarding the dynamics of *R*_0_(*t*) and its stochasticity in terms of the linear power of the estimation error and provide information on how such stochastic behavior is correlated to the outcome of the ongoing pandemics.

**Figure 1.**
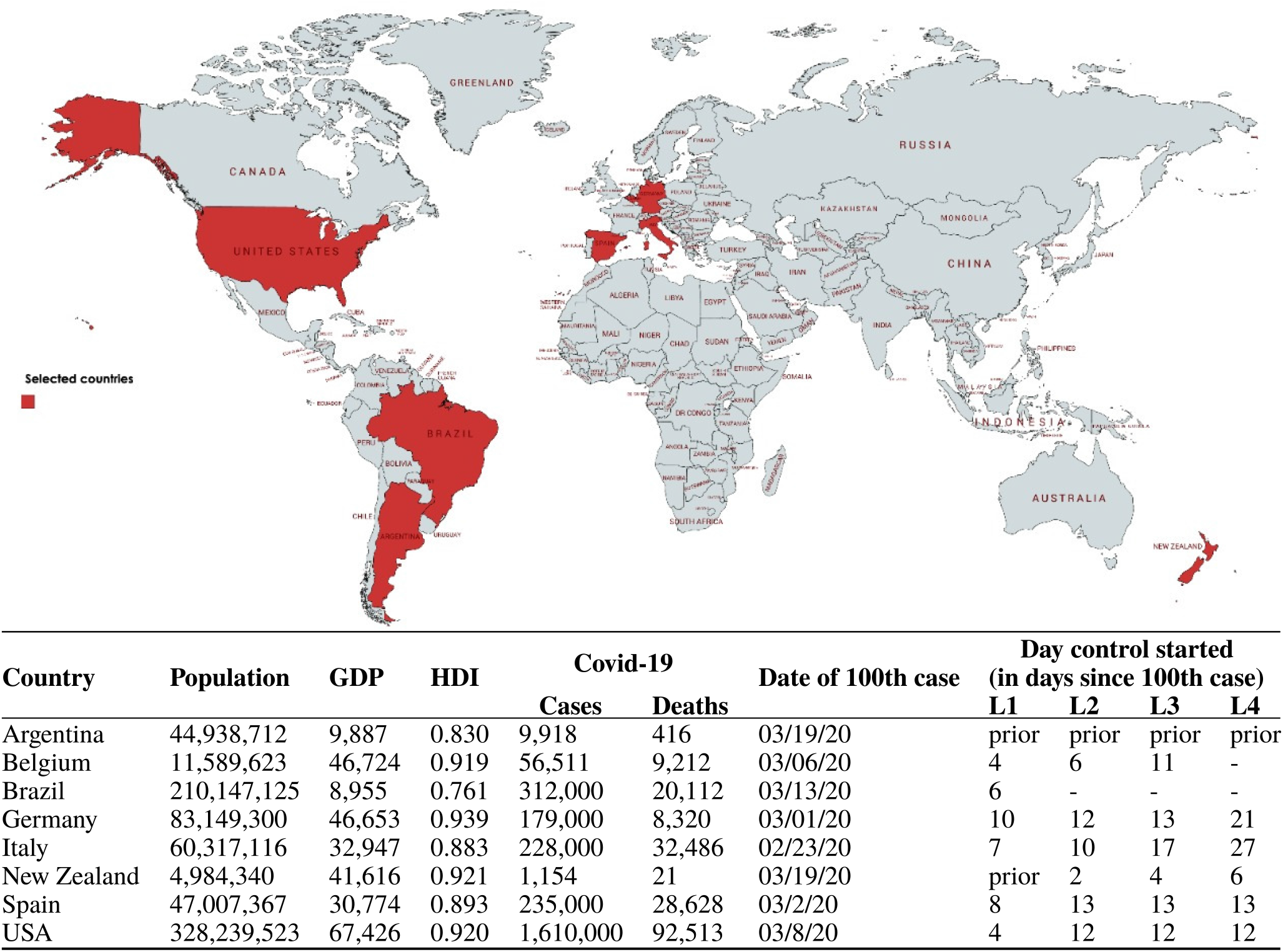
Sociodemographic data for the eight countries targeted in this study and associated COVID-19 data. COVID-19 data accessed on 22/05/20 on https://coronavirus.jhu.edu/map.html. Intervention data according to [40]. GDP per capita in US$. Interventions are categorised into Alert Levels 1 to 4 according to the New Zealand framework (L1 = Level 1, Prepare; L2 = Level 2, Reduce; L3 = Level 3, Restrict; L4 =Level 4, Eliminate) [40].

The LSA is one of the most popular estimation methods in machine-learning and has been used in many scientific and engineering applications [29, 30, 31], including epidemiology, for calibrating mathematical models’ parameters based on time series data while also generating disease forecasts in the near or long terms [14, 32, 33, 34]. While it has been used for centuries as a classic curve-fitting technique [30, 31], it is still a basic tool in modern data science because of its least-squares Euclidean *ℓ*_2_-norm minimization that is advantageous over other norms and metrics, such as the *ℓ_∞_* and *ℓ*_1_ norms, granting reduced sensibility to outliers due to the squared error [35, 31]. Higher-order norms and the use of more generalized cost functions, both linear and non-linear, often require gradient descent solutions which are the foundation of deep learning algorithms with none, many, or infinite solutions. In comparison, LSA is computationally inexpensive and can even be solved analytically.

LSA estimation is based on the least mean-squares algorithm (LMS), a special case of Bayesian estimation and the foundation of classical optimal estimation theory, where its applicability is commonly attributed to offline *batch processing*, i.e., the whole data set must be *a priori* available and be processed at once “in one single step into the estimate” [35], involving complex algebraic procedures such as the inversion of high order matrices, with dimension equal to the length of the data set. Its offshoot, the recursive least-squares algorithm (RLS), has been used for real-time estimation applications in diverse areas such as signal and data processing, communications, and control systems [36, 37, 38], since it benefits from the recursive method and avoids matrix inversion by working one sample at a time, both speeding up processing and avoiding a possible ill conditioned (non-invertible) information matrix formation.

Some of the advantages of RLS over LMS, and other more complex gradient descent-based estimation methods, are: its recursive or sequential processing, which requires less memory over a single iteration step; the possibility to capture the dynamics of non-linear and time-varying systems; its native discrete-time synthesis to deal with discrete-time signals; and its dead-beat convergence characteristics, i.e., convergence in minimal time [30]. However, one important drawback is the possibility of RLS getting stuck in local minima and becoming unstable when tracking certain classes of signals, as remarked in [30]. Thankfully, the exponential functions used in epidemic modeling are within the stable scope of convergence of the RLS method.

In the present work, we compare the performance of both RLS and LMS on estimating *R*_0_(*t*), in terms of processing speed and accuracy. While both are LSA-based methods apparently differing just by the sequential-recursive and the batch-processing forms of implementation, we are dealing with a general case problem of estimating a random variable *R*_0_ (t) given random variables as well in a maximum-likelihood estimation problem that “implies ignorance of any statistics of the estimated variable” [35], *R*_0_(*t*). Thus, if the available measurements are independent, RLS and LMS should achieve the same accuracy performance. Otherwise, the RLS should perform better.

Our modeling approach is based on a discrete-time MIMO setup which uses a well established concept in the design of aerospace navigation systems called sensor fusion and which is summarized by the following statement “one always gains by adding a new measurement in terms of navigation error, and this no matter how bad the additional measurement is” [39, Remark 4.8]. In the present work, we combine data from the number of susceptibles, infections, recoveries, deaths, and individual parameters of three coupled differential equations in order to improve the estimation of *R*_0_(*t*).

## 2 METHODS

### 2.1 Data sources

The COVID-19 data used in this report is publicly available from The Center for Systems Science and l Engineering of the Johns Hopkins University (JSU CCSE) [41], which maintains a Repository on Github [42].

### 2.2 Procedures

The transmission dynamics of the COVID-19 outbreak is usually described by a compartmental model (SEIR), where (S) susceptible – (E) exposed – (I) infected – (R) removed [43, 44]. In a closed population of *P_n_* individuals, the transitions between the compartments (cf. Fig. 2) are described through the following set of differential equations:

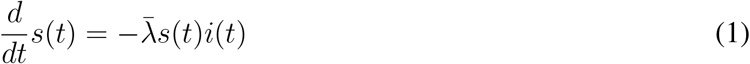

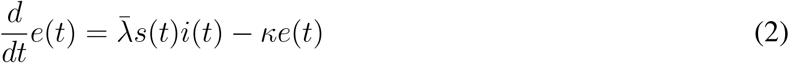

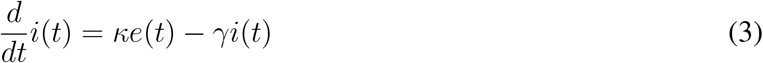

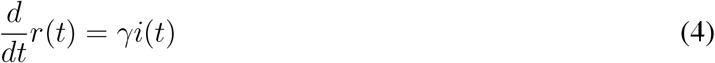

where 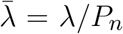, *λ* is the infection rate, *γ* is the remove rate, and *κ* is the incubation rate. From these parameters, it is possible to calculate the basic reproduction ratio, *R*_0_ = *λ*/*γ*. Thus, *R*_0_ is not solely dependent on the infection rate, but also on the frequency of removals due to death or recoveries.

**Figure 2.**
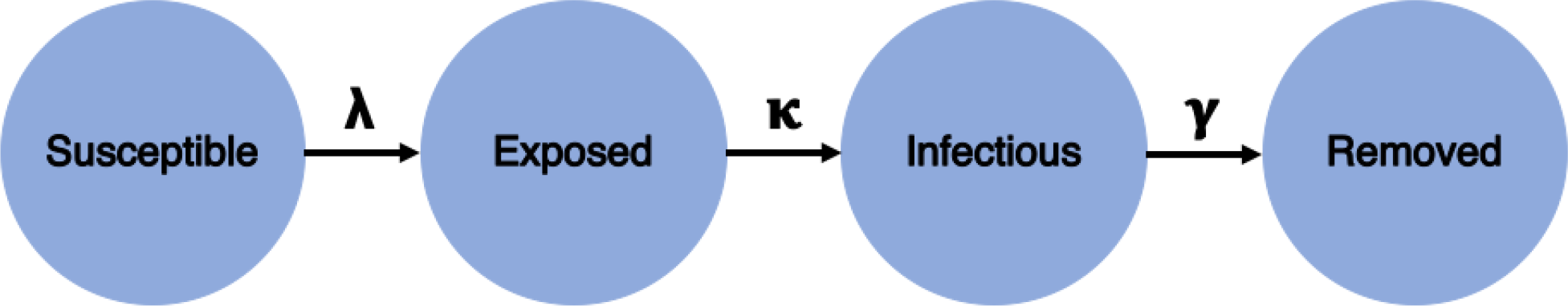
SEIR Model Structure

Assuming that the incubation period of the disease is instantaneous and the duration of infectivity is the same as the length of the disease, we can consider both groups *E* and *I* as contagious and *E*(*t*) := *I*(*t*). Also, according to the Akaike information criterion (AIC), the simpler SIR model performs much better than an SEIR model in representing the information contained in the confirmed-case data available for COVID-19 [45]. The basic SIR model is described by the set of Kermack-McKendrick equations [46]:

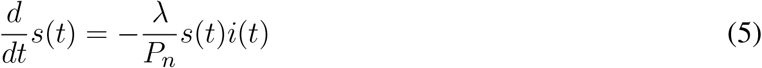

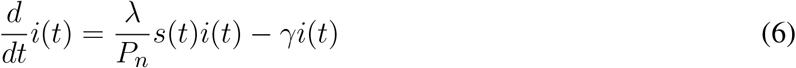

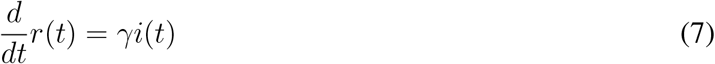

### 2.3 Discrete-time SIR system parametric estimation in real-time

Traditionally, mathematical epidemiology models have been approached with a continuous-time perspective, due in part to the fact that these are more tractable mathematically [47]. However, in order to use machine learning techniques there is a need for a discrete-time equivalent realization to cope with) daily-sampled data [48]. Due to the slow dynamics of the pandemics, a first order continuous to discrete Euler approximation can be applied to the Kermack–McKendrick equations.

For a general *f*(*t*) function, a Backward discrete-time derivative approximation is:

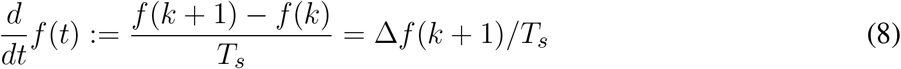

where *T_s_* = 1 is the sampling interval in days and Δ = 1 − *q*^−1^ is the discrete difference operator, defined in *q*−^1^, the backward shift operator domain. The discrete-time approximations of equations (5) to (7) are given by the following difference equations, respectively:

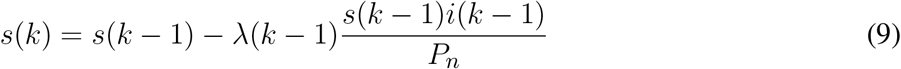

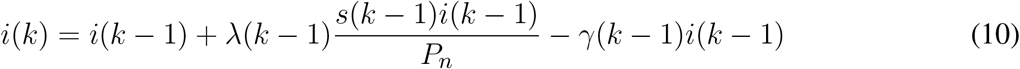

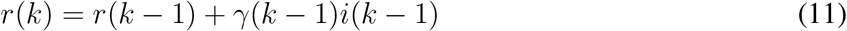

The discrete-time SIR system described above considers time-varying parameters in order to continuously adapt the model as new data becomes available. Using the time-series of infections and removals (due to death or recovery), equations (9) to (11) can be used to estimate the model parameters.

Since Eq. (11) has an exclusive dependence with *γ*(*κ*), this poses a direct estimation problem which can be stated as “for *N* registered samples, minimize the following quadratic cost function:”

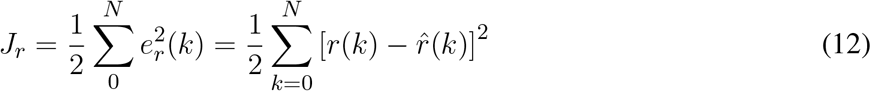

Eq. (12) is based on the estimation error of *r*(*k*). By applying the recursive least-squares method to minimize *J_r_*, it is possible to optimally estimate 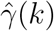 using the following equation:

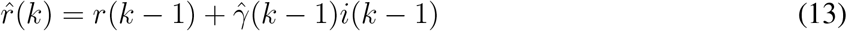

We assume that the estimation error is Gaussian, *e_r_*(*k*) ~ (0, 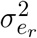), with zero mean and variance 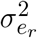, such that *r*(*k*) = *ȓ*(*k*) + *e_r_*(*k*). The estimator gain, the parametric estimation and error covariance minimization are solved recursively, as follows:

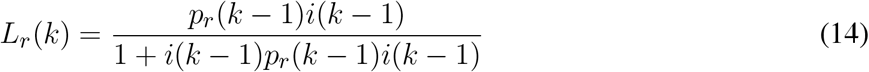

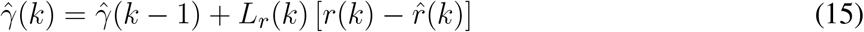

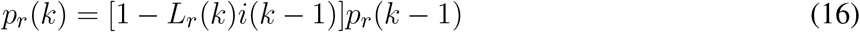

where the error covariance matrix *p_r_*(*k*) can be reset periodically to prioritize more recent data. Specifically in this work, *p_r_*(*k*) is a scalar, since only a single parameter is being estimated. Choices to initialize the covariance matrix may vary, depending on prior available covariance information or other positive definite matrix. The higher its magnitude the higher the estimator gain in the transitory dynamical stage of the estimation procedure.

In regard to Eq. (11) and the estimation of 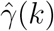, since the infection numbers increase before any removal report is available during the first stages of the pandemic, during this period 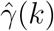 tends to zero and eventually makes the time-varying estimated reproduction number tend to infinity:

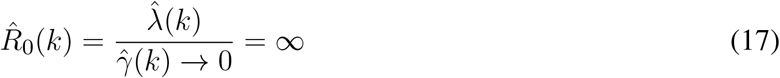

As a consequence of (17), the estimation method proposed in this work cannot be applied when the number of recovered is not available.

Since Eq. (10) depends on both SIR parameters, due to its stronger dependence of 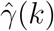 and Eq. (11), its estimated value is substituted into Eq. (10) and another recursive least-squares problem is constructed to estimate 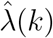, using

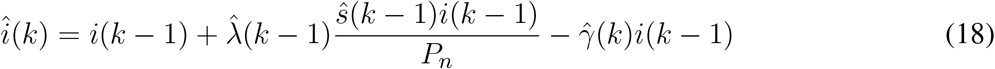

The solution is akin to minimizing the estimation error 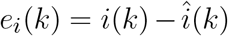, using the following equations for estimator gain, parametric estimation update, and error covariance minimization, respectively:

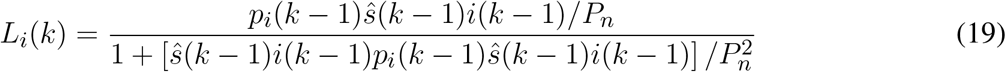

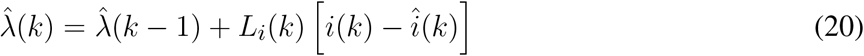

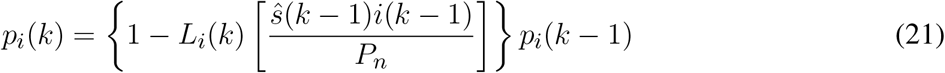

Time-series data for Eq. (9) is not available and the evolution of the Susceptible compartment in time is in fact estimated based on the known initial condition (i.e., the population *P_n_*) and on the estimated 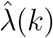. Thus, it is always estimated and fed back to (18), such that the correct form to represent it, within this time-varying SIR model, is by rewriting Eq. (9) based on the estimated Susceptible:

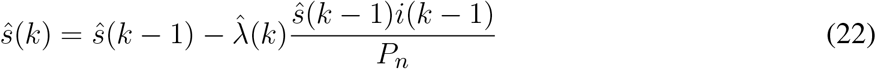

The estimation of the time-varying reproduction number, based on the derived discrete-time SIR model, is given by:

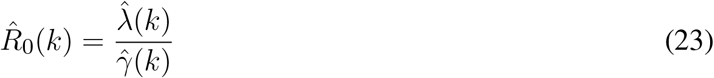

We also adopted two modifications to the nominal *Ȓ*_0_(*k*) equation: a moving 4-day average to compensate ‘ for the randomness of daily updates on incidence data, as seen in the German Daily Situation Report of the Robert Koch Institute on COVID-19 [49], and the proportion of susceptible individuals in the population, l known as the effective reproduction number [14]:

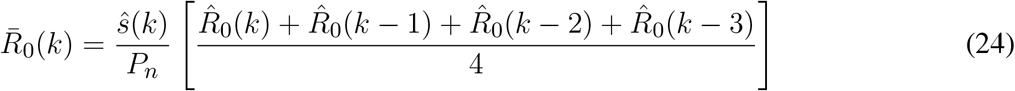

Figure 3 shows a block diagram of the proposed 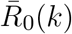 estimator. This diagram presents a clearer view of the coupled multivariate dynamics and closed-loop characteristics of the proposed estimation approach.

**Figure 3.**
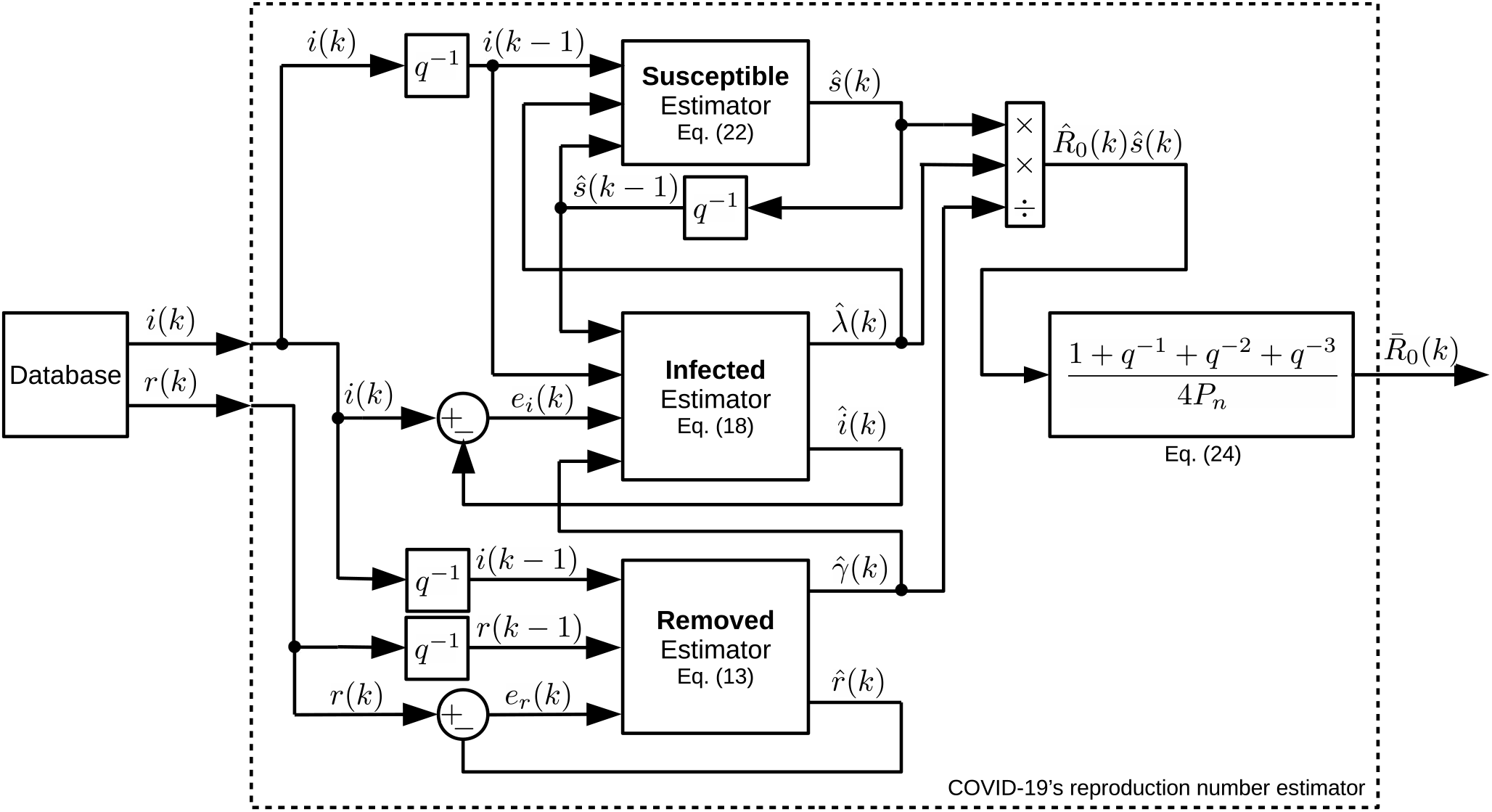
Block diagram of the estimation system. Notice how its subsystems are interconnected using closed-loop strategies and data fusion.

With this formulation, it is possible to analyze the transmission ratio of the pandemics on a daily basis, as with a sensor. Besides, estimations of the transmission ratio produce a dynamic representation from the perspective of the time-series of 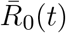 (henceforth designated simply as 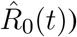), which allows the modeling of its dynamics and its randomness in order to assess stochastic properties correlated to the time-varying ¡ reproduction number, which might reflect how health authorities have been handling the challenges posed by the pandemics in each country considered in this work.

### 2.4 Modeling of *R*_0_

Henceforth we assume that we are able to estimate the reproduction number on a daily basis and it is thus possible to consider it as another output of the proposed pandemic model. Thus, relying on the time-series of *Ȓ*_0_(*k*) and knowing it is correlated with the number of infected and removed individuals, we deploy machine learning techniques to identify a dynamic system that fits the data.

Differently from the real-time monitoring/sensing procedure adopted to estimate *Ȓ*_0_(*k*), now we are interested in obtaining a general model that can describe the dynamics of a system, *Ȓ*_0_(*q*^−1^), for a certain period of interest. In order to model such a system, we used the LMS, which is a non-recursive least-squares estimation technique [31, 30], and propose a black-box linearized polynomial model:

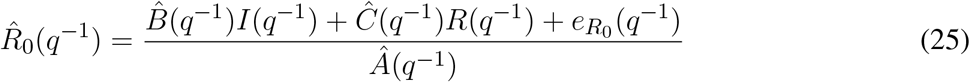

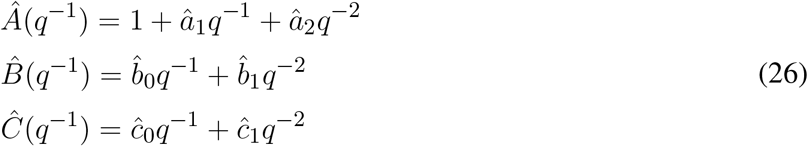

where 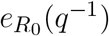 is the Gaussian process based on the estimation error 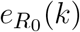 of estimated polynomials shown in (26).

This second-order autoregressive with exogenous inputs (ARX) based model structure is assumed considering the fundamental simplicity of the SIR model, in which the Infected and Removed systems together form a second-order system.

The non-recursive least-squares estimator is a batch processing technique, used to optimally estimate the set of parameters that minimizes a quadratic performance index as the one shown in (12), but using a vector-matrix form of error, 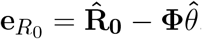. This vector-matrix system is defined as:

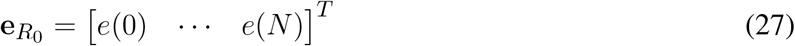

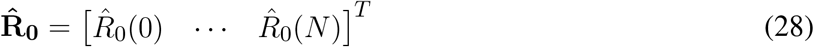

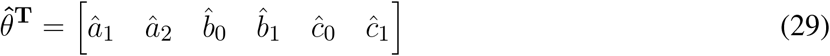

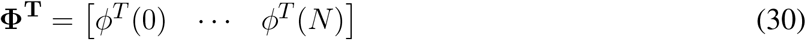

The above equations represent, respectively, the vector of errors, the vector of observed outputs, the estimated parameters vector and the matrix of regressors. The latter is based on the vectors of regressors formed up to *N* registered samples, with such vectors defined as:

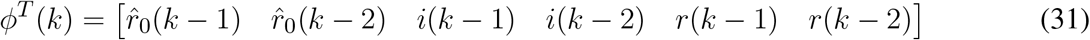

The solution to obtain the estimated parameters is straightforward and given by

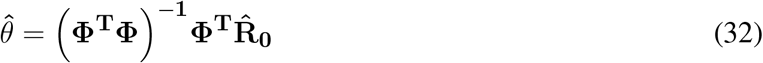

By assuming an ARX linear model it is possible to evaluate the pandemics from the perspective of linear stochastic systems theory, assessing how the reproduction number decays linearly in time in different countries and how the random nature of events associated with the pandemics affects the model’s uncertainties, i.e., 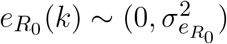. This calculated uncertainty can give us some clues regarding the effectiveness of pandemic control measures.

We propose that by analyzing the linear power of the *Ȓ*_0_(*q*^−1^) estimation error, i.e. 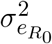, we can use this stochastic property to generate *stochastic ratio* curves based on the estimated linearized model of Eq. (25). The higher the variability associated with the stochastic ratio the higher we expect the variance or linear power of the error to be and, consequently, the more uncertain is the official COVID-19 data reported by health authorities.

## 3 RESULTS

We used publicly available data to validate the algorithms and the estimated time-varying SIR model parameters. This section is organized in the following way: we first present the estimated results based on the number of infectious and removals available from different countries. Then, we present time-series of daily estimates on *R*_0_(*k*) based on the RLS-estimated 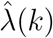 and 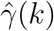 and compare both processing time and accuracy results to the classical LMS method to verify the efficiency of the proposed technique.

These results are followed by the presentation of linearized estimated outputs generated by the ARX-based *Ȓ*(*q*^−1^) models (shown in (25)) using non-recursive or batch processing of 30-days of *R*_0_(*k*) estimates. The modeling residuals were assumed to be Gaussian (zero mean). Estimated error variances were used to produce 200 discrete Gaussian sequences as surrogates to additive white noises, depicted by 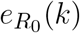 in (25). These 200 additive noise sequences were used to generate, for every analyzed country, 200 stochastic reproduction ratio trajectories, shown together with the linearized ratio and the real-time estimated ratio. It must be remarked, however, that this value of 200 Monte Carlo simulations were selected heuristically in order to give the necessary visual information in our figures, such that the readers could verify by themselves how the modeling uncertainties of the pandemics could mislead our judgment of the probable effective reproduction number of COVID-19 during the studied period. The number of Monte Carlo simulations thus affects only a post-modeling phase and do not interfere with the estimation of the reproduction number.

### 3.1 Discrete-time SIR model estimation results

The Achilles’ heel of the RLS algorithm for parametric estimation may be the setup of initial conditions and whether they are optimal or not. However, this is a major problem only if the estimated parameters are to be used in a real-time adaptive control system where the closed-loop stability must be guaranteed [30]. This is not the case in the present work which is interested only in the modeling question itself. Thus, the initial RLS parameters can be either arbitrarily set or set at zero since, theoretically, the RLS estimator is dead-beat and converges in the minimum possible number of iterations [31, 35].

The discrete SIR model proposed in this work is described by three coupled differential equations, each based on a single recursive regression and thus forming a third order system. Then, theoretically, as a dead-beat estimator, the RLS would take three iterations to converge and estimate optimal parameters. However, we used plenty more iterations than the theoretical requirements, by commencing to process data from 22-March-2020 onwards but evaluating the results just after 23-April-2020, thus giving more than 30 days/iterations for the RLS to converge to an optimal set of parameters by 23-April-2020, when we started our analysis.

The estimators were implemented with arbitrary initial parameters of 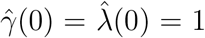 and the initial guess for *Ȓ*_0_(0) = 1. The magnitudes of the estimation error covariance matrices were initialized as *p_r_*(0) = *p_i_*(0) = 1, considering that the initial error is large. Both *p_r_*(*k*) and *p_i_*(*k*) were reset to 1 every 7 days to prioritize more recent data [50]. The selection of the reset period considered not only the number of days required to wash-out outliers but also to allow weekly changes in the parameters’ dynamical behavior, such as due to lockdowns or quarantine relaxation. we also adopted a moving 4-day average for *Ȓ*_0_(*k*), shown in (24), to compensate for daily random effects [49]. We only used data from 22 March 2020 onwards, when all eight countries already had more than 100 infections reported.

Figures 4 and 5 show the dynamics of both Infected and Removed cases using the SIR model. It must be remarked that the Infected curve in the United States was downscaled by a factor of 4 in order to fit in the graph along with the other studied countries. It is evident from Fig. 4 that the RLS provide parametric estimates that fit the reported data. The same goodness of fit cannot be observed in Fig. 5, as the number of removals for the U.S., for example, poses some difficulties for the RLS estimator, as depicted by the estimated removals (dotted lines) showing a certain dispersion from the real data (continuous lines). However, the inclusion of the Removed data set, even with bad measurements, leads to a better estimate of *Ȓ*_0_(*k*).

**Figure 4.**
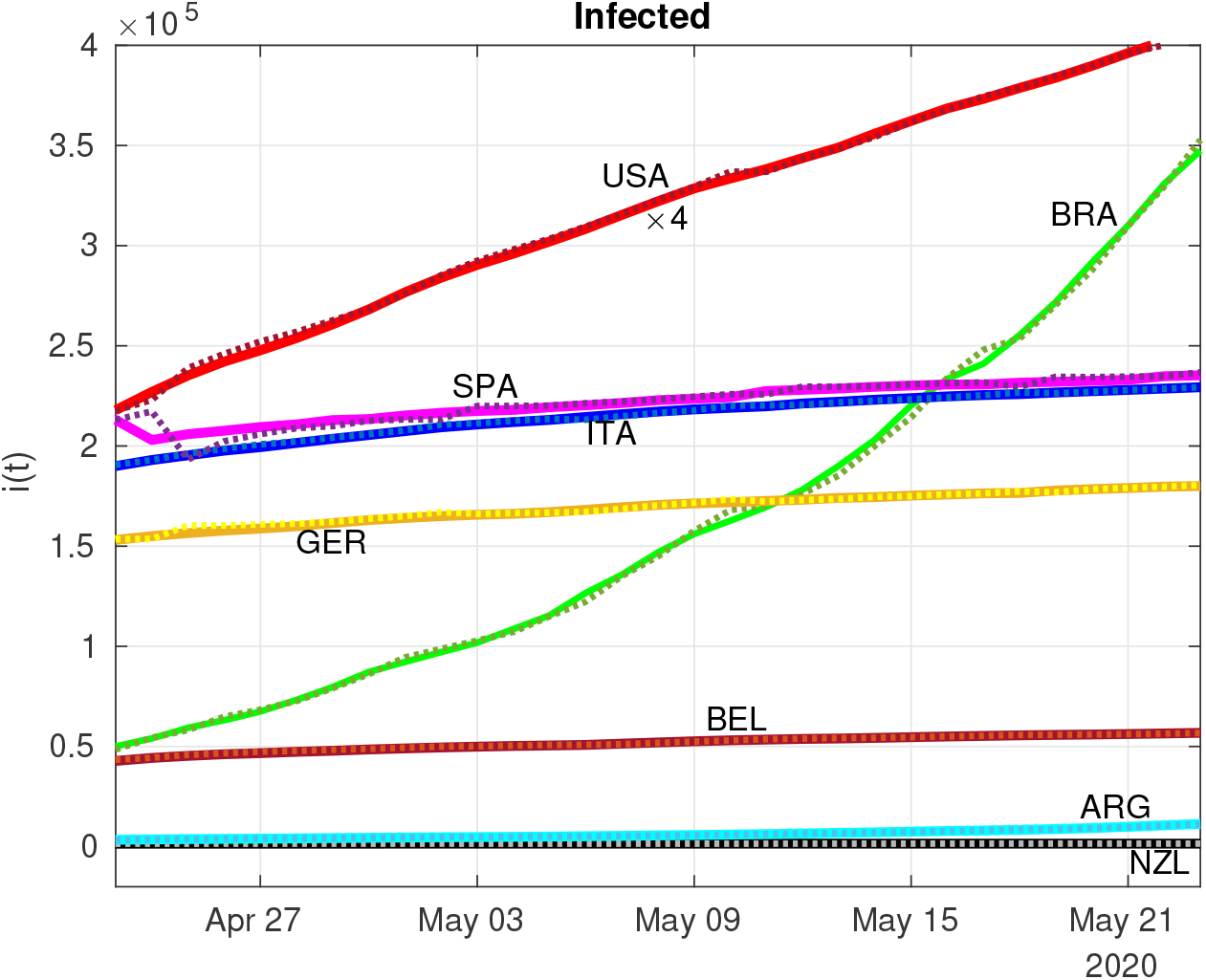
Estimation (dotted lines) of Infected individuals (continuous lines) using the RLS technique (The USA data was downscaled 4 times to fit in the graph).

**Figure 5.**
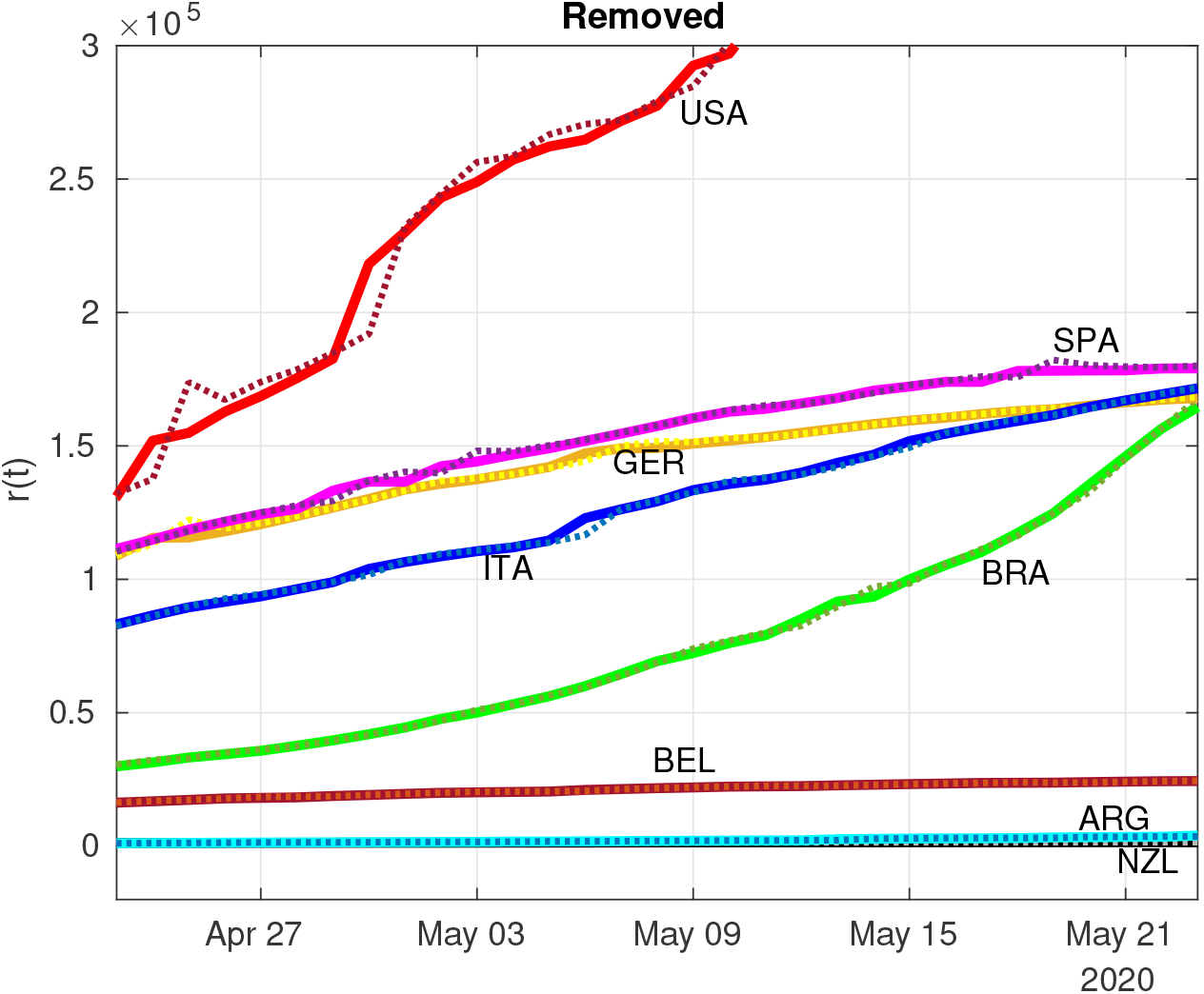
Estimation (dotted lines) of Removed individuals (continuous lines) using the RLS technique.

Fig. 6 presents the *Ȓ*_0_(*k*) of the eight different countries during a period of two weeks. Despite the large variability of both Argentina and the United States, there are other countries that are already reducing the number of new infections and where the frequency of removals has increased, such as in Germany, Italy,) and New Zealand (see Table 1).

**Figure 6.**
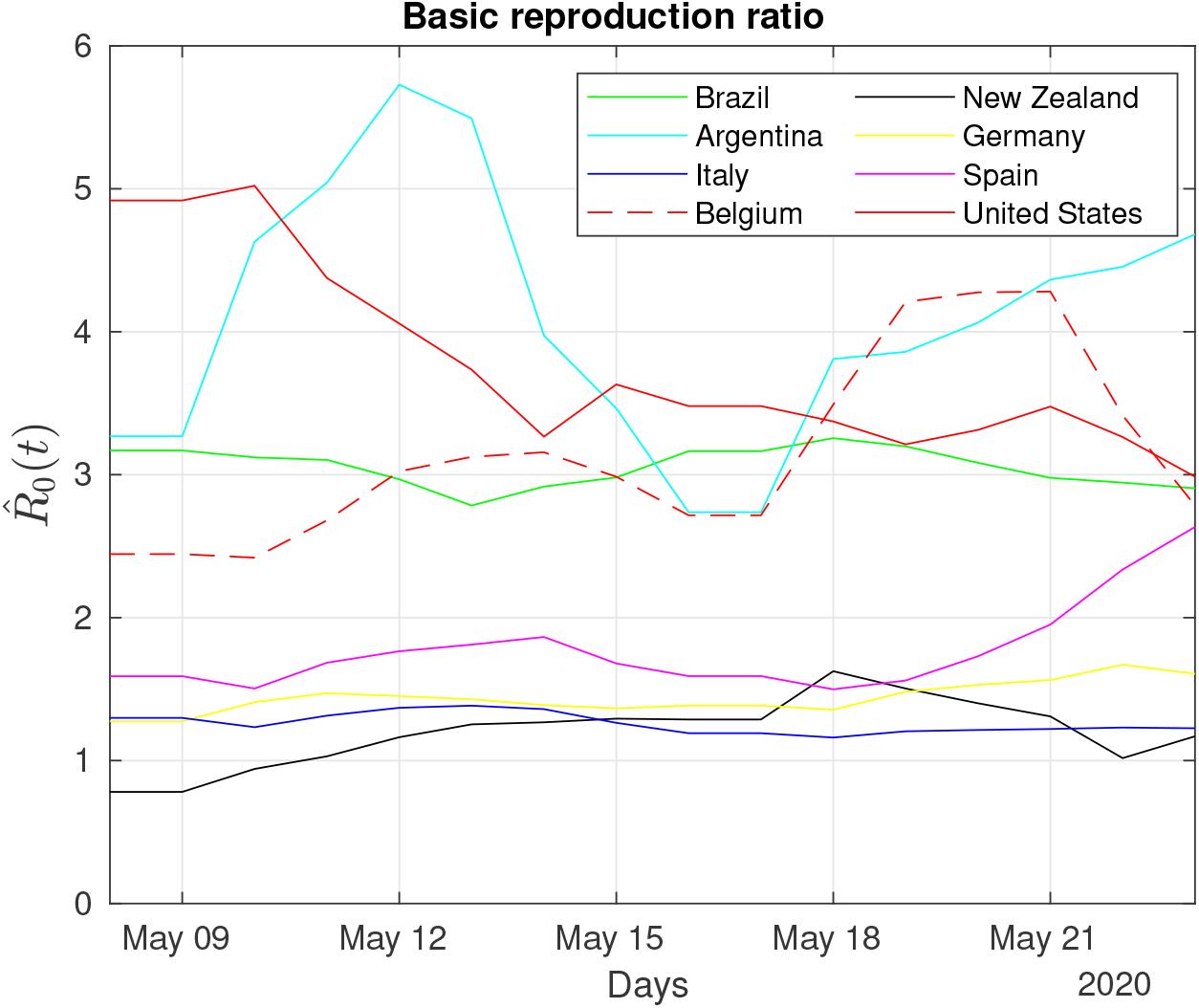
Daily monitoring of the COVID-19 pandemic: dynamics of the estimated reproduction number for two weeks in May, according to available official data and the SIR model.

**Table 1.** Recoveries (May 21, 2020).

| Country | Recoveries |
| --- | --- |
| Argentina | 30.5% |
| Belgium | 26.6% |
| Brazil | 40.5% |
| Germany | 88.2% |
| Italy | 59.0% |
| New Zealand | 96.6% |
| Spain | 70.3% |
| United States | 23.5% |

The trajectory of the curves shown in Fig. 6 can also be associated with some extraordinary events that occurred during the same period. For example, after 3 May 2020, when some U.S. states had relaxed social distancing guidelines, it is possible to observe a corresponding phasic increase in the basic reproduction ratio of the U.S., which was also reported in the Washington Post on 9 May 2020 [51].

One interesting trajectory in Fig. 6 regards Argentina. For most of the time, the estimated reproduction ratio of Argentina was one of the highest and comparable only to the U.S., despite its low number of infected individuals (cf. Fig. 4). This apparent contradiction is related to the low number of removals at the beginning of the pandemic that tends to raise the *Ȓ*_0_(*k*) (see Eq. 17). Recoveries in Argentina, based on data of May 16 (cf. Table 1), are approximately 30.5% of its total infected, close to Belgium with 26.6%) and whose transmission ratio is below Argentina’s, reinforcing the notion that the transmission ratio is not a static parameter and is best approached by dynamical systems theory.

The results displayed in Figure 4 seem at odds with a recent report that claimed that Brazil’s *R*_0_ had recently dropped to 1.4 [52]. However, on the same day a Brazilian newspaper quoted this report, the country had a record number of new infections and deaths [53]. This reinforces the notion that machine learning methods might give better and faster clues regarding the severity of the pandemic in terms of *Ȓ*_0_(*k*).

We compared the performance of the proposed RLS estimator with its most common counterpart, the LMS algorithm. In this comparison, we considered the average processing time and the normalized relative estimation error based on the mean-square-based cost function shown in Eq. (12). The average processing time results for the RLS and LMS were, respectively, 14 ms and 20 ms. We used a computer with a 4th Generation Intel Core i5-4200U CPU at 1.6 GHz, 4 GB of RAM, running Ubuntu 18.04 LTS and Matlab R2018a (Mathworks, Inc.). The accuracy results expressed by normalized relative errors are summarized in Fig. 7. Notice that the RLS outperformed the LMS in all cases, except for Argentina.

**Figure 7.**
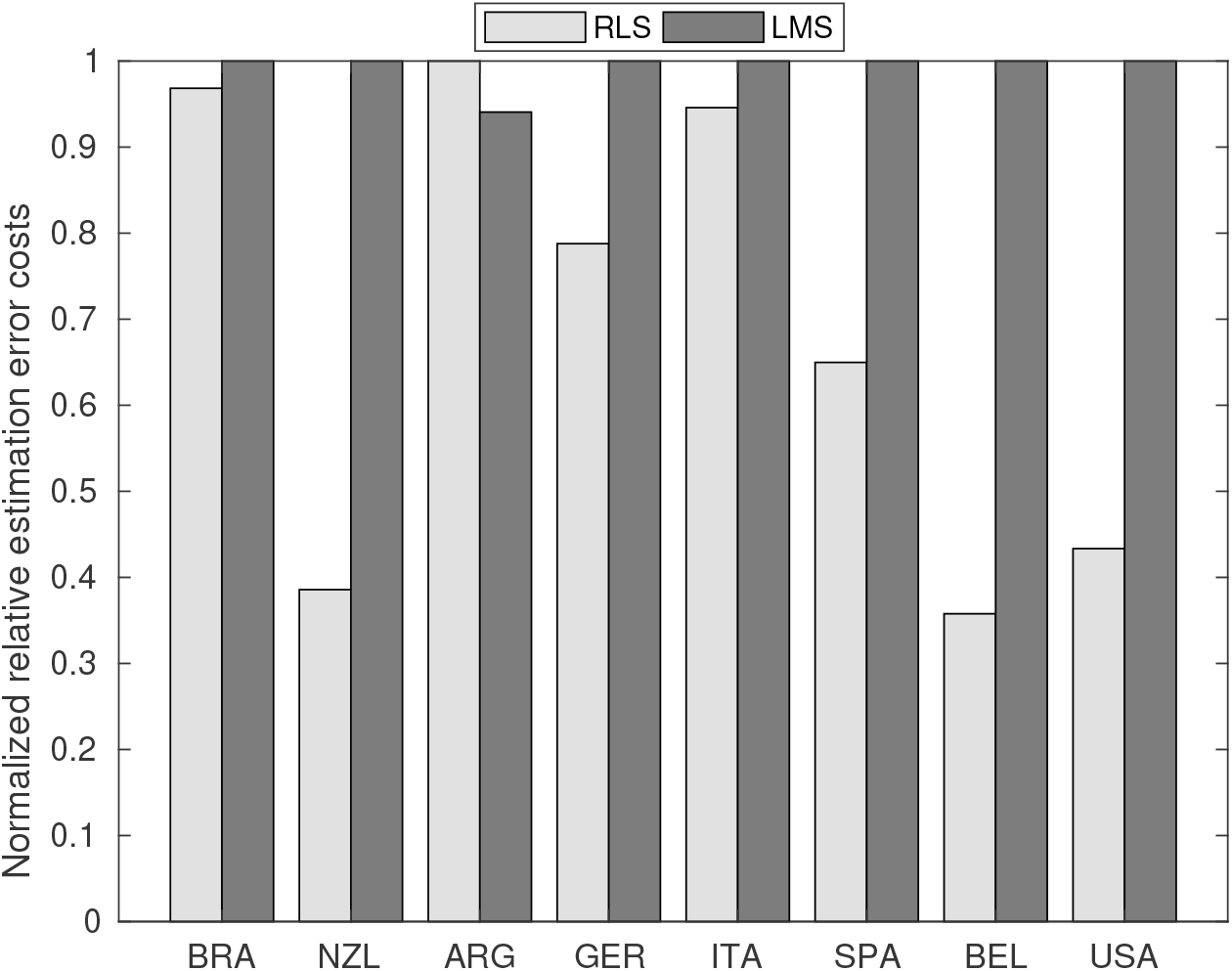
Comparison of normalized relative estimation error costs between the proposed RLS-based technique and the traditional LMS estimation algorithm. The average processing time of RLS and LMS was, respectively, 14ms and 20ms.

### 3.2 Assessing the pandemics through *R*_0_ dynamics

Figures 8 to 11 show *Ȓ*_0_(*k*) for a 30-day period for the eight investigated countries. These figures show linearized *R*_0_ together with the respective real-time estimated data that originated this second stage estimation. The variance of the estimation error is then used to generate Gaussian sequences which are superimposed on the linearized *R*_0_ estimate, the stochastic ratio which synthetically reproduces the stochasticity and uncertainties of *R*_0_ estimates. Both Germany and Italy (Fig. 8) are through a period of consistently decaying *R*_0_(*t*). Despite the hard way COVID-19 hit Italy before, its stochastic ratio currently is among the lowest.

**Figure 8.**
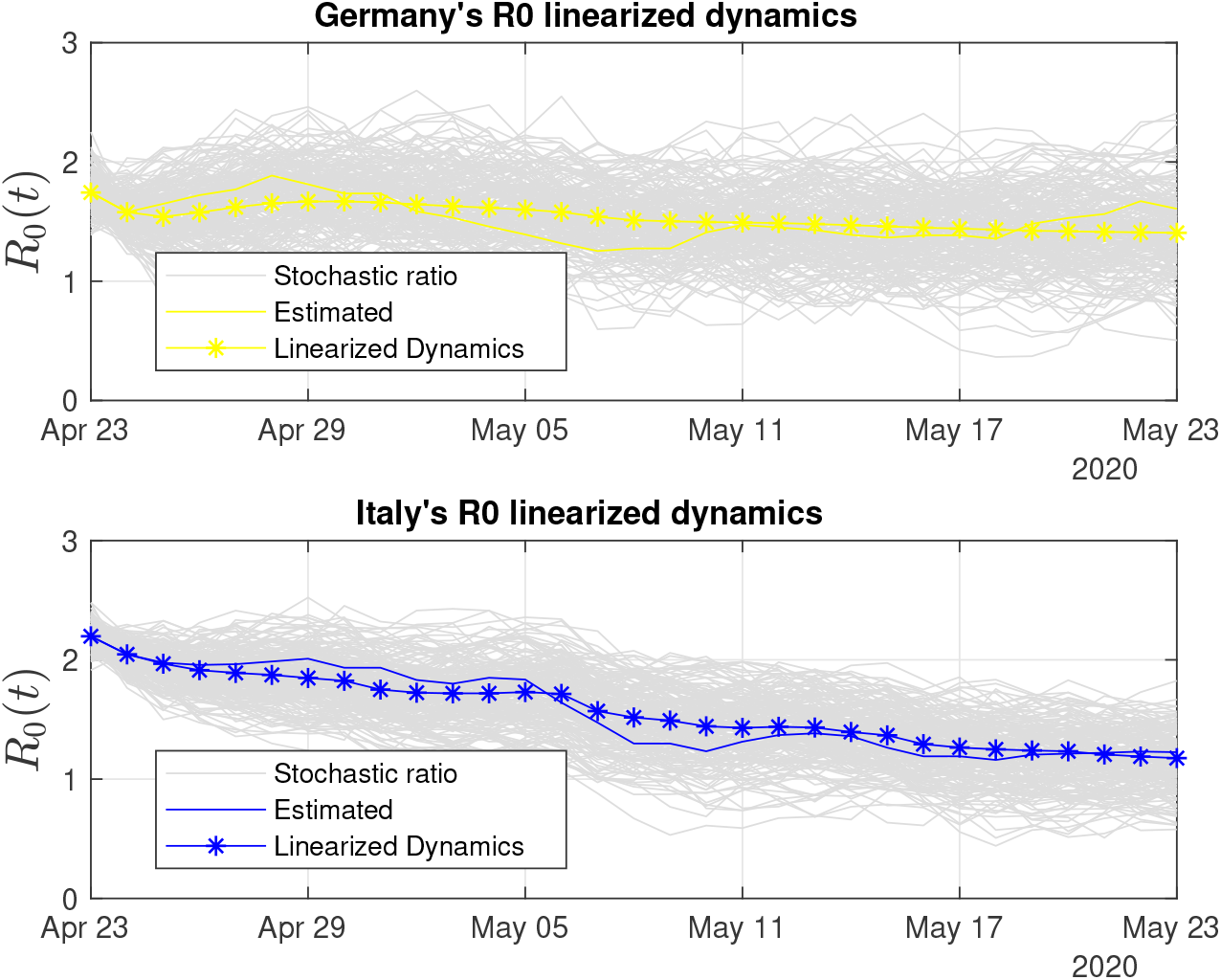
30-days of linearized R0 dynamics for Germany and Italy. Both countries have low variability in the stochastic ratio curve.

**Figure 9.**
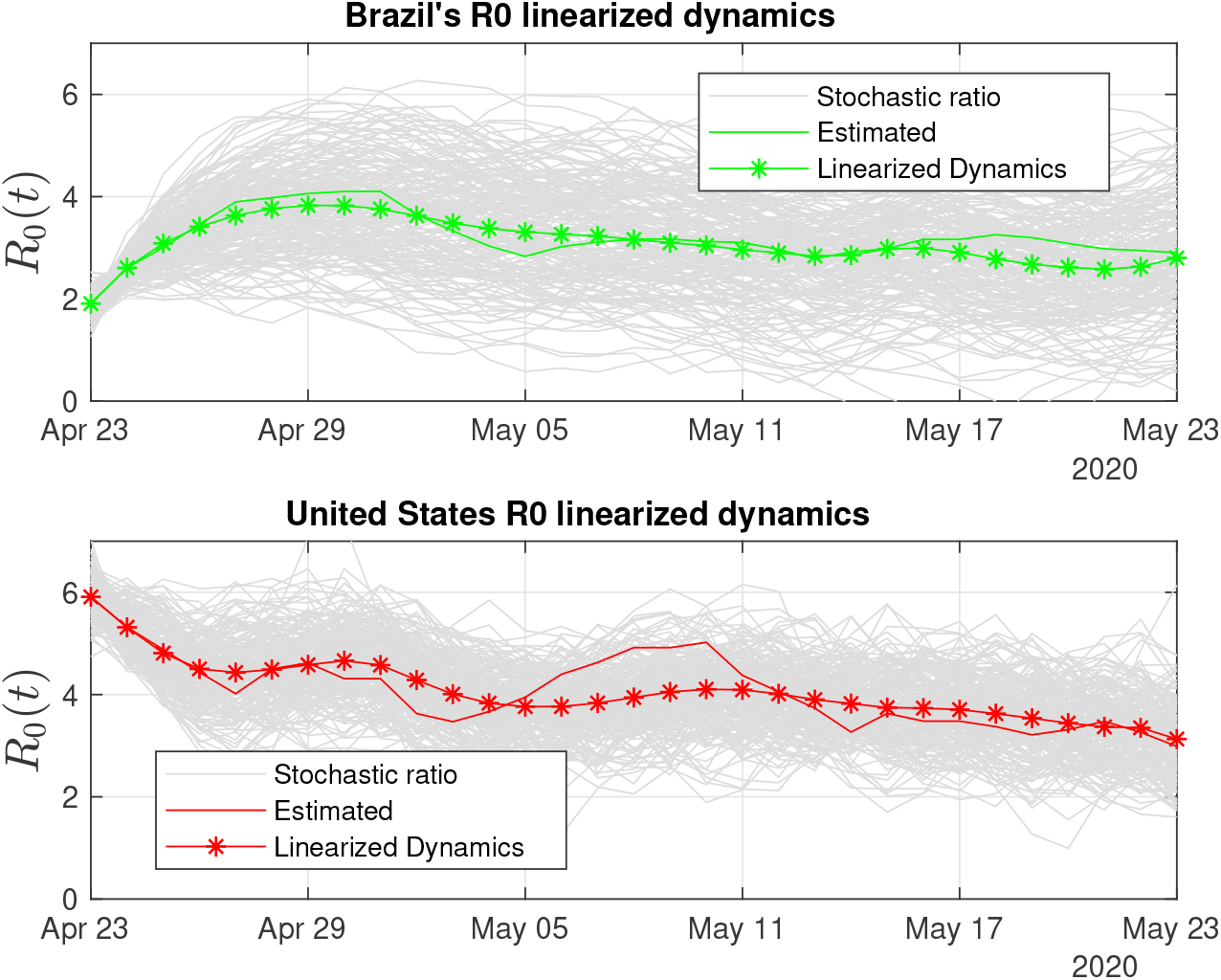
Brazil and the US display large variability in the stochastic ratio curve, which reflects data uncertainty and difficulties to control COVID-19’s spread.

**Figure 10.**
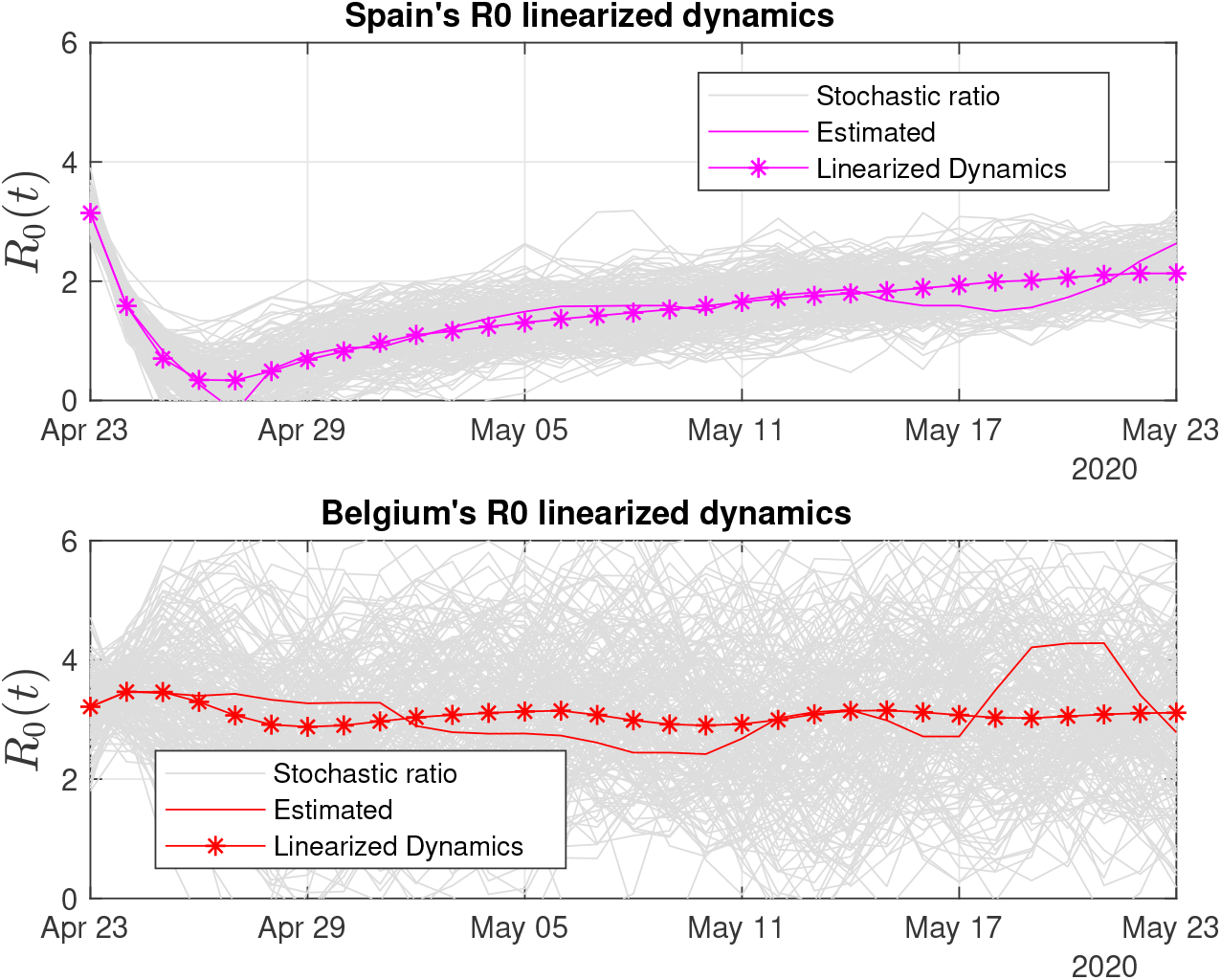
30-days of linearized *R*_0_ dynamics for Spain and Belgium. Both countries exhibit high variability of the stochastic ratio, which is suggestive of problems with either data compilation or difficulties to stabilize the pandemics.

**Figure 11.**
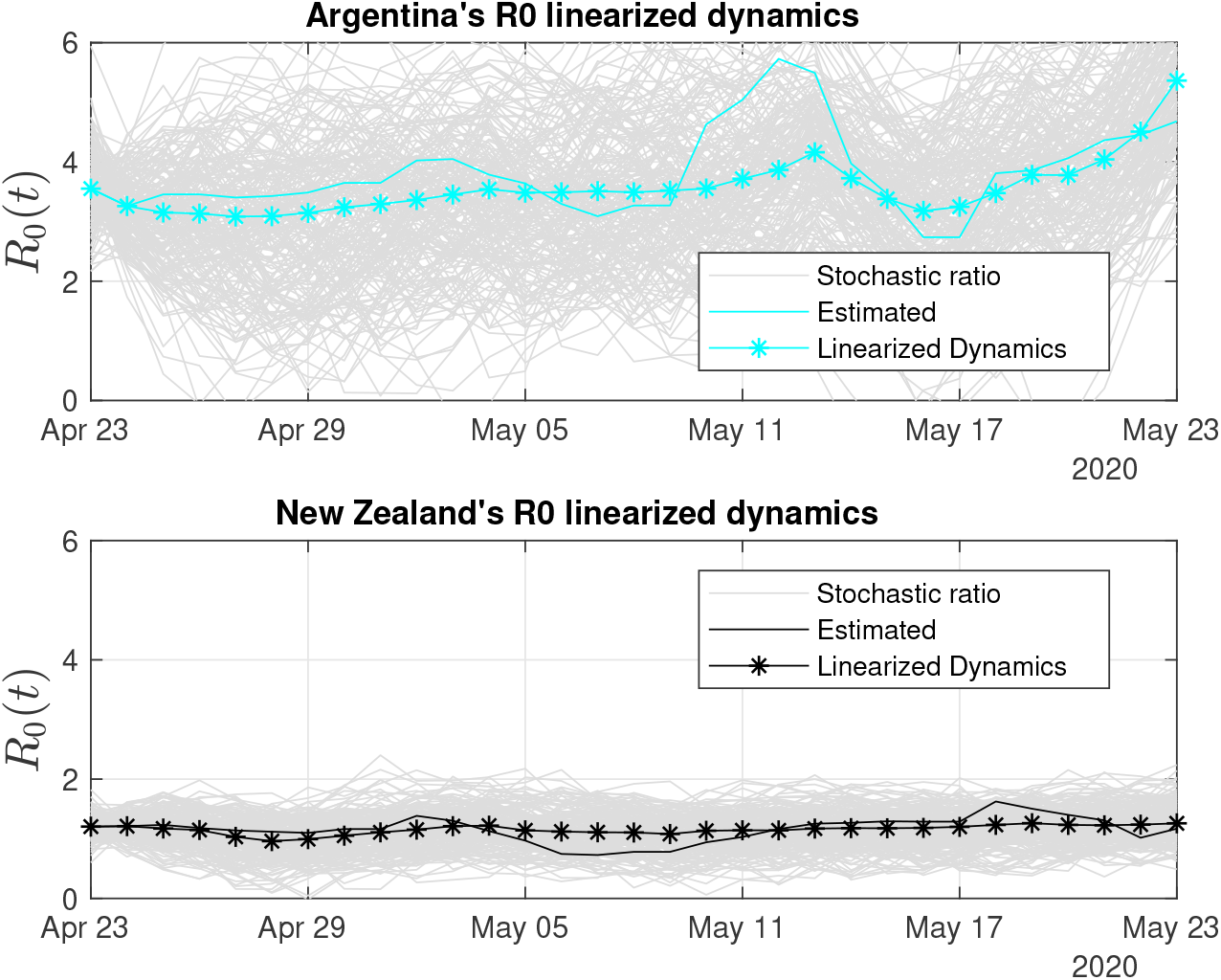
30-days of linearized *R*_0_ dynamics for Argentina and New Zealand. New Zealand has been able to control the infection rates, which is reflected on the low variability stochastic ratio curve.

Brazil and the United States (Fig. 9) are the two most populous countries of our sample and the most hard-hit by COVID-19 among them. Both countries also have been struggling with their uneven response to the pandemics [54]. This outcome is captured by our *R*_0_ sensors, with the Brazilian stochastic ratio being in decrease, as Recovered data becomes more available (see Fig. 9).

In Fig. 10 we present linearized *R*_0_ estimates for Spain and Belgium. Spain’s estimates have suffered the influence of annotation errors (by subtracting infected individuals in 24 Apr. 2020) which eventually provoked oscillations in the real-time ratio estimate, making it zero-cross on April 27th. Such an error was washed out by the linearized estimate and has certainly contributed to its increased degree of uncertainty (see Table 2).

**Table 2.** Pandemic’s uncertainty (up to 23 May 2020).

| Country | Power | Uncertainty |
| --- | --- | --- |
| Argentina | 0.3099 | 30.9% |
| Belgium | 0.2426 | 24.2% |
| Brazil | 0.0546 | 5.4% |
| Germany | 0.0220 | 2.2% |
| Italy | 0.0113 | 1.1% |
| New Zealand | 0.0349 | 3.4% |
| Spain | 0.0521 | 5.2% |
| United States | 0.1835 | 18.3% |

Belgium’s degree of uncertainty was the worst among all the countries, at least during the period we analyzed. One possible clue to understand why Belgium’s stochastic ratio became so variable is to consider the impossibility to linearize its dynamics and the associated increase in error. However, our estimation procedure considers the error as Gaussian, and its mean value in fact approaches zero. Thus, the linearized estimate based on 30-days of data has a high probability to be close to the values shown.

Belgium has also shown an increased *R*_0_ during the analyzed period. The low number of recoveries (see Table 1) influenced the frequency of removals and the basic reproduction number, which is the ratio between the frequency of infections and the frequency of removals (see Eq. (23)). Even though a recent report [55] proposed that Belgium’s *R*_0_ at the beginning of May 2020 was 0.8, according to our adaptive SIR-based *R*_0_ estimations, the current situation in Belgium is uncertain with a probable *R*_0_ close to 3.0, 320 which is the mean value of 30-days linearized dynamics (see Table 3).

**Table 3.** R0 estimates (on/up to 23 May 2020).

| Country | Real-time | Linearized | 30-days Mean |
| --- | --- | --- | --- |
| Argentina | 4.6 | 5.3 | 3.5 |
| Belgium | 2.7 | 3.1 | 3.0 |
| Brazil | 2.9 | 2.8 | 3.0 |
| Germany | 1.6 | 1.4 | 1.5 |
| Italy | 1.2 | 1.1 | 1.5 |
| New Zealand | 1.1 | 1.2 | 1.1 |
| Spain | 2.6 | 2.1 | 1.4 |
| United States | 2.9 | 3.1 | 4.0 |

Figure 11 shows the trends for Argentina and New Zealand. The *R*_0_ estimates for the two countries, which have the lowest number of infections of the group, suggests that several recent *R*_0_ reports are considering a ratio solely based on the number of infections and population, without estimating and taking into account the frequency of removals, which makes *R*_0_ increase.

We compiled a table of the uncertainty of the estimated stochastic ratios by country (see Table 2), where the percentage of uncertainty is proportional to the linear power of the Gaussian estimation error. Table 3 shows the estimated *R*_0_ estimates by country and demonstrate the usefulness of our approach to provide a real-time picture of the pandemic that can be used to support decision-making.

## 4 DISCUSSION

Almost two months after the WHO declared COVID-19 a global pandemic, health complications due to SARS-CoV-2 have caused many deaths and upended the routines of billions of people around the world. Research efforts for the development of a vaccine are being accelerated but it is still a distant target [56]. At this moment, however, the most effective interventions are NPIs aimed at reducing SARS-CoV-2 transmission rates in order to increase the fraction of severe cases having access to scarce medical resources, such as mechanical ventilation [57].

Mathematical modeling is a valuable instrument to gauge the epidemics’ dynamics and evaluate the effects of interventions aimed to control its spread. A crucial parameter is *R*_0_, the basic reproduction number, which is closely followed by health officials and the public alike. As the world hopefully transitions to a gradual release from social distancing measures, many questions still remain about the SARS-CoV-2 virus and there is all but the inevitability of secondary waves of infection ahead. Thus, the continuing use of mathematical models to track the disease will remain a necessity. However, the utility of models depends on the quality of the data they are fed and there are many uncertainties regarding publicly available data on COVID-19 cases. For instance, due to the lack of widespread testing and sub notifications on the cause of deaths due to the disease, it is almost impossible to have a definitive picture of transitions between the compartments used to model the disease. Thus, in this work, we provide a system that takes into consideration the degree of uncertainty of the results presented by epidemiological SIR-type models.

The NPIs aimed to control the spread of the the COVID-19 pandemic are implemented in a way similar to feedback control systems, with health authorities implementing social restriction measures in response to real-time evaluation of the number of infected and removed cases. Therefore, as in control systems, we are interfering with COVID-19’s dynamics, affecting its behavior and parameters, and even discussing the development of real-time monitoring techniques using automation and control systems technologies. However, such closed-loop control systems still need humans in the loop and some data annotation mistakes may occur such as: seasonal dynamic changes related to workers shifts, weekend reduced reports and even possible data manipulation of the data log may compromise the stability of estimators.

We showed that RLS performed a better estimation job than LMS in the present work, both in speed and accuracy. Real-time parametric estimation techniques, such as RLS, are widely used in the automation and aerospace industries to support adaptive control systems and state estimators. In automation applications, the RLS and LMS estimators are in fact divided into two different technologies, respectively, self-tuning and auto-tuning [30], where the first refers to real-time online adaptive process and the second involves the following steps: on-demand user request, acquiring the data set with sufficient samples, run offline processing without time constraints, present the results to the user decide whether to deploy or not the parametric update. Auto-tuning is less costlier to implement and thus more common, whereas the self-tuning technology requires online real-time processing with time constraints and is more commonly applied in the aerospace industry [58, 30].

LSA-based estimation is becoming more popular in industrial settings, such as: in single-input, single-output systems (SISO) and in multiple-inputs, multiple-outputs systems (MIMO); in linear and non-linear system identification, and in polynomial and state-space MIMO system realizations [59, 60]. Such an increase in the use of RLS for real-time applications is justified by the advances of microprocessors to cope with the time constraints of fast dynamical systems, where the sampling interval can go as short as a few nanoseconds. Modern engineering applications with great impact in our society are associated with discrete-time sampled systems and signals, thus requiring adequate and reliable digital estimation algorithms such as LSA. At least from an engineering viewpoint, the COVID-19’s pandemic is also a discrete-time system, since the publicly available data for infections, deaths and recoveries are updated on a daily basis, thus generating discrete-time sequences with the sampling period of 24h.

During COVID-19’s pandemic the scientific community is struggling to study, design and deploy engineering applications to better assist society with information that could somehow quantify the pandemic’s degree of severity based on *R*_0_(*t*) estimates [61]. Different from traditional modelling approaches, we propose a discrete-time MIMO system realization instead of the continuous-time estimation using ordinary differential equations; the MIMO-based parametric estimation of *R*_0_(*t*) is obtained through data fusion in a closed-loop estimation arrangement (see Fig. 3).

The innovation of our modeling approach is the use of a discrete-time MIMO setup based on sensor fusion, a well established strategy employed in the design of aerospace navigation systems under the assumption that “one always gains by adding a new measurement in terms of navigation error, and this no matter how bad the additional measurement is” [39, Remark 4.8]. Thus, by fusing data from the number of susceptibles, infections, recoveries, deaths, and individual parameters of the three coupled dynamical equations instead of a single one based solely on infections, the chance of having a better estimate of *R*_0_(*t*) in the maximum-likelihood sense is higher. Of course, the drawback of our proposed approach is the impossibility to apply it to nations lacking publicly reports on death and recovery numbers.

Our comparative approach shows that the strict measures adopted by some countries managed to stabilize the epidemics, such as Germany, Italy, Spain, and New Zealand. In others, such as the U.S. and Brazil, the delay in adopting such measures and lack of coordination proved decisive to keep *R*_0_ values high and with a high degree of uncertainty. The real-time estimation of model parameters such as *R*_0_ provides important insight into the underlying epidemic process and provides robustness in the face of imperfect data. This strategy can be eventually implemented as an online tracking application providing information about the dynamics of the pandemic to health officials and the public at large.

## Data Availability

Data referred in the manuscript is available upon request to the authors

## CONFLICT OF INTEREST STATEMENT

The authors declare that the research was conducted in the absence of any commercial or financial relationships that could be construed as a potential conflict of interest.

## AUTHOR CONTRIBUTIONS

A.S. and A.P. conceived and wrote the manuscript.

## FUNDING

Antonio Silveira acknowledges the financial support of the Brazilian Research Agency, the CNPq (*Conselho Nacional de Desenvolvimento Científico e Tecnológico*), under grant 408559/2016-0.

## DATA AVAILABILITY STATEMENT

The datasets analyzed for this study can be found in the COVID-19 data repository from The Center for Systems Science and Engineering of the Johns Hopkins University (JSU CCSE) [41, 42].

## Notes

### Competing Interest Statement

The authors have declared no competing interest.

### Clinical Trial

The study used publicly available data from internet databases

### Funding Statement

No specific funding was received for this work

